# Diagnosing Chronic Obstructive Airway Disease: a diagnostic accuracy study of a smartphone delivered algorithm combining patient-reported symptoms and cough analysis for use in acute care consultations

**DOI:** 10.1101/2020.09.05.20164731

**Authors:** Paul Porter, Joanna Brisbane, Udantha Abeyratne, Natasha Bear, Javan Wood, Vesa Peltonen, Phill Della, Fiona Purdie, Claire Smith, Scott Claxton

**Affiliations:** School of Nursing, Midwifery and Paramedicine, Curtin University, Bentley, Western Australia, 6102; PHI research group, Joondalup Health Campus, Joondalup, Western Australia, 6027; School of Information Technology and Electrical Engineering, University of Queensland, Brisbane, Queensland, Australia, 4072; Institute of Health Research, University of Notre Dame, Western Australia, 6959; ResApp Health, Brisbane, Queensland, Australia, 4000; Genesis Care Sleep and Respiratory, Perth, Western Australia, 6027

## Abstract

**Background:** Rapid and accurate diagnosis of Chronic Obstructive Pulmonary Disease (COPD) is problematic in acute-care settings, particularly in the presence of infective comorbidities.

**Objective:** The aim of this study was to develop a rapid, smartphone-based algorithm for the detection of COPD, in the presence or absence of acute respiratory infection, and then evaluate diagnostic accuracy on an independent validation set.

**Methods:** Subjects aged 40-75 years with or without symptoms of respiratory disease who had no chronic respiratory condition apart from COPD, chronic bronchitis or emphysema, were recruited into the study. The algorithm analysed 5 cough sounds and 4 patient-reported clinical symptoms, providing a diagnosis in less than 1 minute. Clinical diagnoses were determined by a specialist physician using all available case notes, including spirometry where available.

**Results:** The algorithm demonstrated high percent agreement (PA) with clinical diagnosis for COPD in the total cohort (n=252, Positive PA=93.8%, Negative PA=77.0%, AUC=0.95); in subjects with pneumonia or infective exacerbations of COPD (n=117, PPA=86.7%, NPA=80.5%, AUC=0.93) and in subjects without an infective comorbidity (n=135, PPA=100.0%, NPA=74.0%, AUC=0.97.) In those who had their COPD confirmed by spirometry (n=229), PPA = 100.0% and NPA = 77.0%, AUC=0.97.

**Conclusions:** The algorithm demonstrates high agreement with clinical diagnosis and rapidly detects COPD in subjects presenting with or without other infective lung illnesses. The algorithm can be installed on a smartphone to provide bedside diagnosis of COPD in acute care settings, inform treatment regimens and identify those at increased risk of mortality due to seasonal or other respiratory ailments.

## INTRODUCTION

Chronic obstructive pulmonary disease (COPD) is the fourth leading cause of mortality, affecting more than 384 million individuals worldwide [1]. It is characterised by airflow limitation and a progressive decline in lung function [2]. The population prevalence of COPD, via spirometry screening, is reported to be 9-26% in those greater than 40 years old [3]. It is estimated that 80% of people with COPD are undiagnosed [4] and up to 60% of those with a diagnosis of COPD were found to be misdiagnosed upon subsequent spirometry [5, 6]. 30-60% of patients who have been diagnosed by a physician with COPD have not undergone spirometry testing [7]. In a study of 533 COPD patients, 15% of those with spirometry tests did not show obstruction and 45% did not fulfil quality criteria [8].

COPD should be considered in patients who present with dyspnoea, chronic cough, sputum production or recurrent lower respiratory tract infections and who have been exposed to tobacco or air pollution. Airflow limitation, demonstrated by a FEV_1_/FVC ratio of < 0.7 on post-bronchodilator spirometry is considered diagnostic of COPD according to criteria stipulated by the Global Initiative for Chronic Obstructive Lung Disease (GOLD) [2]. The severity of airflow limitation in COPD can be classified by the degree of reduction in FEV_1_ as a percentage of the predicted value [2]. However, spirometry is not routinely used in emergency departments or primary care settings due to inexperience, time constraints and availability of equipment [9].

Early and accurate diagnosis of COPD is imperative to ensure initiation of correct treatment, particularly as evidence suggests the incipient stages represent a period of rapid decline in lung function where cessation of smoking and targeted intervention may be of value [10]. Rapid identification and management of COPD is important in acute care settings as there is a heightened risk of mortality from respiratory infections such as seasonal influenza [11]. SARS-CoV-2 has a reported case fatality rate of 1.4% for patients without comorbid conditions vs 8.0% for those with chronic respiratory conditions [12].

Screening for COPD in primary care settings using spirometry in asymptomatic patients has not been found to be efficient as high numbers need to be screened to detect any cases [13]. Screening questionnaires such as the COPD diagnostic questionnaire (CDQ) have performed poorly in an asymptomatic cohort in the primary care setting [14]. We propose that the best use of an algorithm for screening is in a scenario where patients present to a healthcare facility with symptoms, where there is a higher pre-test probability of case detection.

We have previously demonstrated high diagnostic agreement of an automated algorithm with clinical diagnoses for paediatric respiratory diseases including croup, asthma, bronchiolitis and pneumonia. The algorithm also accurately separated upper-from lower-respiratory tract conditions [15]. The technology, which has regulatory approval, is similar to that used in speech recognition software and combines lower airway audio data transmitted during cough events and simple patient-reported clinical symptoms to derive the diagnostic probability output [16]. As the lower airway is open to the outside during a cough, sounds are transmitted through the mouth and can be recorded. In this way, it is similar to traditional auscultation; however, much higher bandwidth is achievable using our method as the chest wall is no longer reducing sound transmission. We recorded audio by a standard smartphone where the in-built diagnostic algorithm provided a rapid result without requiring clinical examination or additional diagnostic tests.

In this paper, we describe the development and evaluate the accuracy of an algorithm for diagnosing COPD from a cohort of mixed respiratory disorders, including acute respiratory infections. The intended use population is those who present to health settings with suspected respiratory illness.

## METHODS

### Study Population and Setting

Between Jan 2016 and March 2019, a convenience study sample was obtained by prospectively recruiting participants from the emergency department, low-acuity ambulatory care and inpatient wards of a large, general hospital in Western Australia; and the consulting rooms of a respiratory physician.

This diagnostic accuracy study is part of a more extensive development program (Breathe Easy / ANZCTR: ACTRN12618001521213). Subjects were approached if they presented to a participating site with signs or symptoms of respiratory disease or to specialist rooms for a lung function test. Subjects with no discernible symptoms of respiratory disease were also recruited. Subjects were excluded if they were on ventilatory support, had a terminal disease, were medically unstable, had structural upper airway disease or had a medical contraindication to providing a voluntary cough (e.g. severe respiratory distress; eye, chest or abdominal surgery within three months; history of pneumothorax). Subjects with uncontrolled heart failure/cardiomyopathy, neuromuscular disease or lobectomy/pneumonectomy were also excluded. From this cohort, only those aged 40-75 years were used for the COPD development program.

Written informed consent was obtained from all participants, and the study was approved by a Human Research Ethics Committee (Reference Number: 1501). There were no adverse events reported. The study did not interfere with clinical care with all treatment decisions were at the discretion of the treating physician.

### Index Test (Software algorithm)

The development of the mathematical techniques used to derive the algorithm has been described in depth elsewhere [15-18]. Briefly, an independent training cohort (n=564) was used to obtain clinical data and cough samples (from which mathematical features were extracted). In developing the algorithm, selected features were weighted and combined to build various continuous classifier models used to determine the probability of a COPD diagnosis (reference test). The probability output of the algorithm represents the specific, weighted combination of features. Multiple clinical symptoms and audio characteristics were examined and combined with the goal to reduce the number of inputs to a minimum and to use patient-reported symptoms rather than clinically determined signs, vital signs or investigations. Each input added to the overall accuracy and discriminatory clinical ability of the algorithm. The optimal model and corresponding probability decision threshold was selected using a Receiver Operating Characteristic (ROC) curve with due consideration given to achieving a balance of PPA and NPA [16]. Once the optimal model was developed, it was locked from further development and prospectively tested for accuracy on an independent testing set.

Audio data was obtained from 5 coughs using a smartphone (iPhone6) held approximately 50cm away from the subject at a 45-degree angle to the direction of the airflow. Recordings were undertaken in standard clinical environments; however, we took care to avoid other people’s coughs and voices. The cough recording was obtained within 30 minutes of the physical examination of the patient to ensure the clinical features had not changed. If the subject was unable to provide 5 coughs that were recognised by the cough-detection software or the cough recording became corrupted, the subject was excluded from further analysis.

The following 4 clinical symptoms were selected for inclusion in the tested model: subject age; smoking pack-years and subject-reported presence of acute cough or fever during this illness. One smoking pack-year was defined as 20 cigarettes or 20 g tobacco, smoked each day over one year [19]. Where the clinical symptoms were partially unknown, the algorithm did not return a response.

### Reference test (Clinical Diagnosis or Spirometry)

A full medical assessment was performed on all participants at the time of enrolment, including history and clinical examination. Diagnostic tests were ordered by the treating clinician independently of the study and results were available to researchers.

A specialist physician assigned a clinical diagnosis to each subject based on a review of their medical file, including discharge diagnosis, all outpatient and inpatient notations and radiology/laboratory results. The same clinical diagnosis definitions (Table 1) were employed in both the testing set (described here) and in the training set used for algorithm development:

**TABLE 1:** Clinical Diagnosis Definitions

|  |  |
| --- | --- |
| COPD | - Respiratory symptoms consistent with COPD and history of smoking (>10 pack-years)/environmental exposure AND:<br>o If spirometry performed, then FEV1/FVC <0.7 on the best test (after bronchodilator) OR<br>o If spirometry not performed, then previous physician-diagnosis of COPD |
| COPD (infectious exacerbation) | - ALL OF:<br>o Met COPD case definition (as above),<br>o Worsening symptoms of shortness of breath (SOB), cough;<br>o Signs and symptoms of acute respiratory tract infection |
| Acute lower respiratory tract infection (LRTI) | - New lower-respiratory tract symptoms (SOB, cough, chest pain (<1 week) and acute fever AND:<br>o For pneumonia: New consolidation on chest x-ray (CXR) or CT, OR<br>o For LRTI: Infiltrate but no consolidation on CXR or CXR not performed. |
| No lower airway disease | - No lung disease and spirometry results within normal parameters (FEV1/FVC > 0.7 on best test). |

Spirometry was performed according to standard methodology [2, 20].

### Analysis population

Diagnostic accuracy tests were performed for four groups using an independent, test set of subjects. The same inclusion and exclusion criteria were used for both training and test sets (Table 2):

**TABLE 2:**
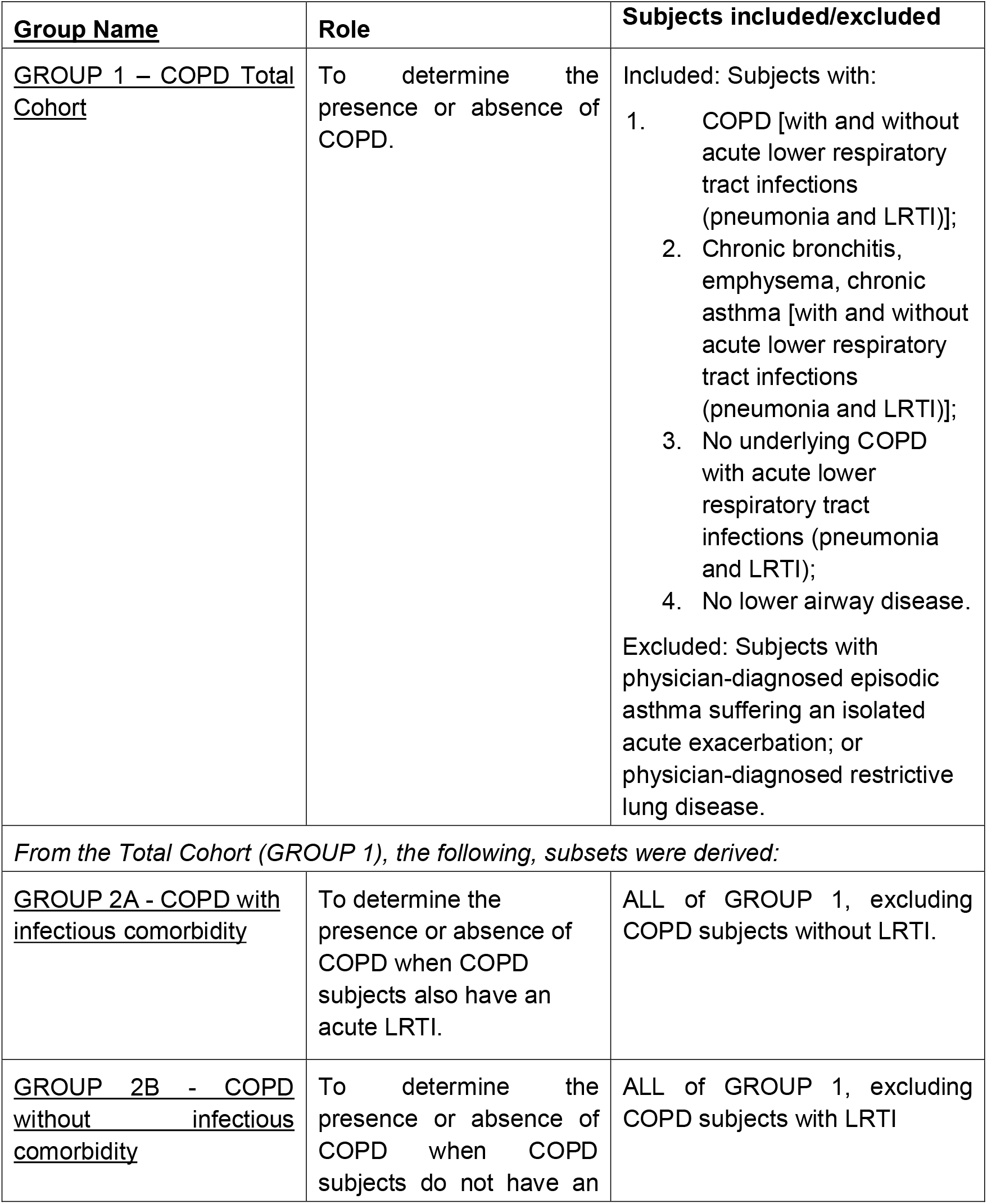
Analysis Groups

After a clinical diagnosis was assigned to all subjects, the database was locked, and the software algorithm was run by an independent researcher to ensure blinding was maintained. Each subject’s cough sound data and clinical diagnosis were only used once in the prospective test.

### Statistical Analysis

Power calculations were derived as follows. Based on expected positive and negative percent agreement greater than 85% from the training program, to obtain a superiority endpoint of 75% (lower bound 95% CI of maximum width ±0.10) a minimum of 48 cases were required.

Positive percent agreement (PPA) is defined as the percentage of subjects with a positive index test result for a specified condition who also have a positive reference standard for the same condition. Negative percent agreement (NPA) is the percentage of subjects who returned negative results for both tests.

The primary study endpoint was defined as PPA and NPA of the index test with the reference standard, with 95% confidence intervals calculated using the method of Clopper-Pearson. The probability of positive clinical diagnosis was calculated for each subject by the final classifier model and was used as the decision thresholds in the derived ROC curve.

## RESULTS

In the prospective testing set, 270 participants met inclusion criteria for, and were enrolled in the COPD diagnostic study. Of these 153 were from the hospital emergency department or inpatient wards, and 117 were respiratory outpatients or from the ambulatory acute care unit.

Two hundred and fifty-two participants provided a valid index and reference test (figure 1). Two were excluded as the clinical diagnosis was recorded as unsure. The mean age of participants was 59.7 ± 9.2 years, 58.7% were female. Those with COPD were older than those without (65.5 vs 57.8 years, p<0.0001), although the sex proportion did not differ with the diagnosis. 85.3% of the entire cohort had at least one of the following respiratory symptoms: acute, chronic or productive cough; fever; rhinorrhoea; SOB; wheeze; or hoarse voice. Subject characteristics are shown in Table 3, including spirometry results where available.

**Figure 1.**
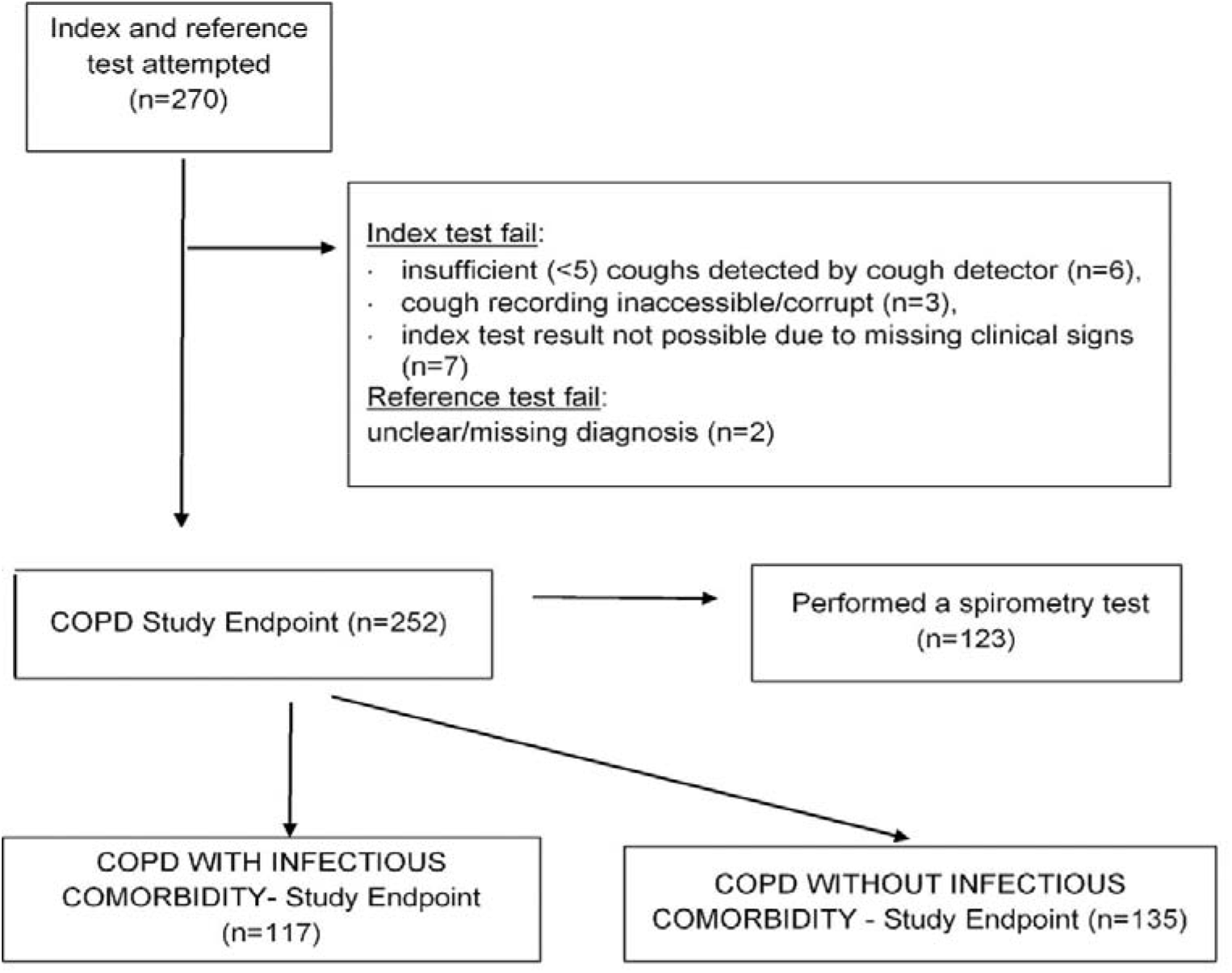
Flow of Participants through the study

**TABLE 3:** Subject characteristics. Data includes all subjects in analysed groups (COPD Positive and Negative).

|  |  | **COPD Total Cohort**<br>**(Group 1, n=252)** | **COPD with infectious comorbidity**<br>**(Group 2A, n=117)** | **COPD without infectious comorbidity**<br>**(Group 2B, n=135)** | **COPD confirmed by spirometry**<br>**(Group 3, n=229)** |
| --- | --- | --- | --- | --- | --- |
| **Age** | Mean (SD) | 59.7 (9.2) | 60.6 (9.1) | 59.0 (9.1) | 59.0 (9.1) |
| **BMI** | Mean (SD) | 28.8 (7.3) | 29.0 (7.9) | 28.6 (6.7) | 29.2 (7.3) |
| **FEV<sub>1</sub>** | Mean (SD) | 2.3 (1.0) | 0.9 (0.2) | 2.3 (1.0) | 2.3 (1.0) |
| **FVC** | Mean (SD) | 3.2 (1.1) | 1.9 (0.4) | 3.3 (1.1) | 3.2 (1.1) |
| **FEV<sub>1</sub>/FVC** | Mean (SD) | 69.1 (16.2) | 46.1 (11.3) | 70.5 (15.4) | 69.1 (16.2) |
| **Predicted FEV<sub>1</sub>** | Mean (SD) | 81.2 (28.8) | 34.3 (12.3) | 84.0 (27.0) | 81.2 (28.8) |
| **Predicted FVC** | Mean (SD) | 90.7 (22.2) | 57.6 (14.0) | 92.7 (21.0) | 90.7 (22.2) |
| **Predicted FEV<sub>1</sub>/FVC** | Mean (SD) | 83.2 (21.9) | 58.4 (14.0) | 85.3 (21.1) | 83.2 (21.9) |
| **Acute cough** | NO | 136 (54.0%) | 22 (18.8%) | 114 (84.4%) | 129 (56.3%) |
|  | YES | 116 (46.0%) | 95 (81.2%) | 21 (15.6%) | 100 (43.7%) |
| **Fever** | NO | 126 (58.1%) | 39 (33.3%) | 87 (87.0%) | 114 (58.8%) |
|  | YES | 91 (41.9%) | 78 (66.7%) | 13 (13.0%) | 80 (41.2%) |
| **Rhinorrhoea** | NO | 116 (53.7%) | 61 (52.1%) | 55 (55.6%) | 101 (52.3%) |
|  | YES | 100 (46.3%) | 56 (47.9%) | 44 (44.4%) | 92 (47.7%) |
| **Wheeze** | NO | 145 (66.8%) | 84 (71.8%) | 61 (61.0%) | 134 (69.1%) |
|  | YES | 72 (33.2%) | 33 (28.2%) | 39 (39.0%) | 60 (30.9%) |

For cases where spirometry (n=123) was used to confirm the presence or absence of COPD, the mean age was 60.0 ± 8.7 years and 65.0% were female with FEV_1_ measurements as shown in Table 4. The COPD negative group includes six chronic, fixed asthmatic patients with FEV_1_ below 80%.

**TABLE 4:** Spirometry derived FEV_1_ (GOLD severity categories) in subjects with and without COPD [8].

| **Percent Predicted FEV<sub>1</sub>**<br>**(GOLD severity category)** | **COPD-Positive n (%)**<br>**of total)** | **COPD-Negative n (%)**<br>**of total)** |
| --- | --- | --- |
| < 30.0%<br>(GOLD 4: Very Severe) | 5 (11.9%) | 0 (0.0%) |
| 30.0% - 49.9%<br>(GOLD 3: Severe) | 17 (40.5%) | 2 (2.5%) |
| 50.0% - 79.9%<br>(GOLD 2: Moderate) | 16 (38.1%) | 4 (4.9%) |
| ≥ 80.0%<br>(GOLD 1: Mild) | 4 (9.5%) | 75 (92.6%) |
| **TOTAL** | **42 (100.0%)** | **81 (100.0%)** |

The calculated PPA and NPA of the algorithm with clinical diagnosis and AUC are in table 5. ROC curves for each test group are in Fig 2.

**TABLE 5:**
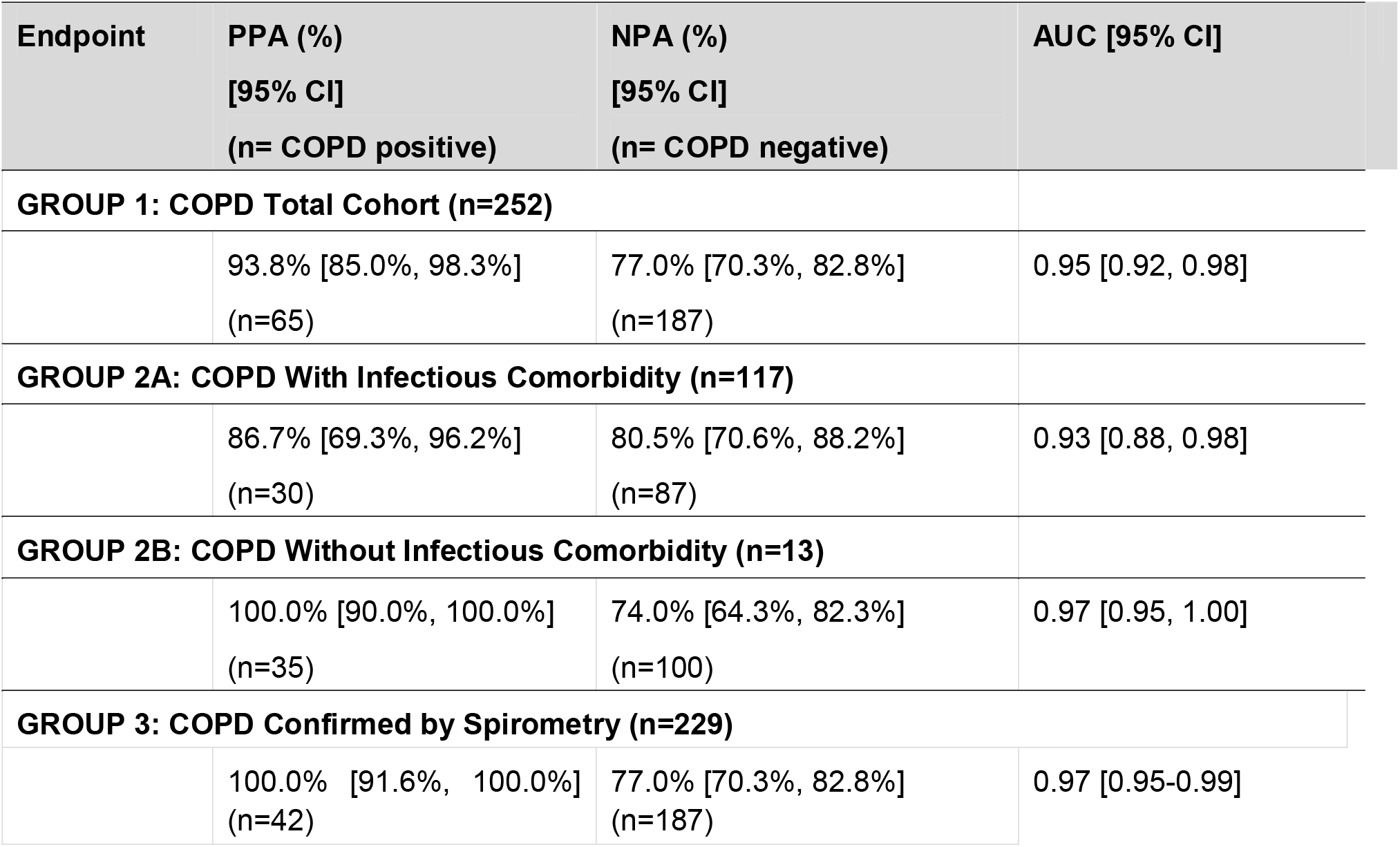
PPA, NPA and calculated AUC of the algorithm (index test) compared with clinical diagnosis (reference test)

**FIGURE 2:**
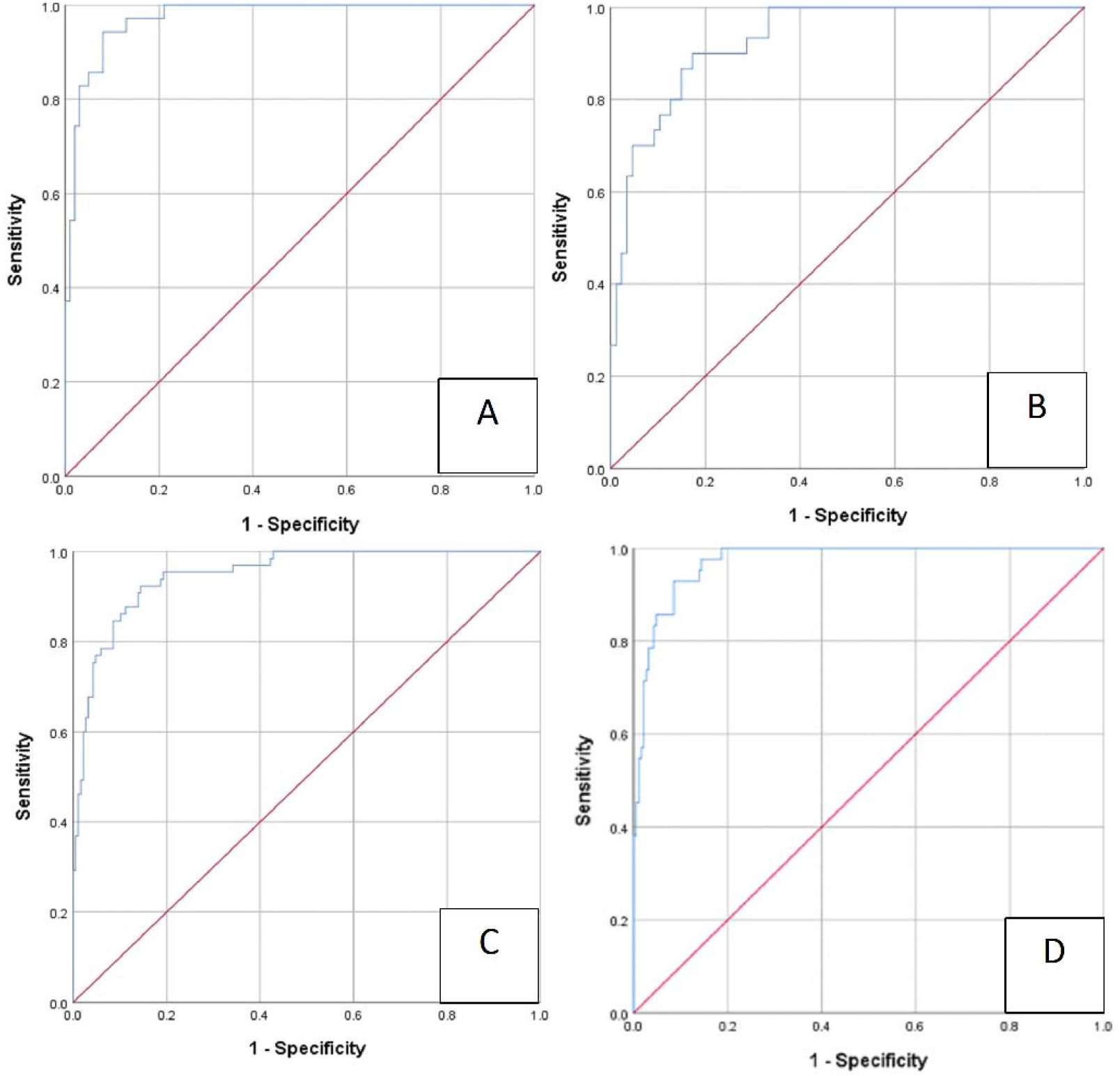
Receiver Operating Characteristic (ROC) Curve and Area under Curve (AUC) for: **A** - COPD Total Cohort (GROUP 1) AUC = 0.95 [95% CI: 0.92, 0.98], **B** - COPD with infectious comorbidity (GROUP 2A) AUC = 0.93 [95% CI: 0.88, 0.98], **C** - COPD without infectious comorbidity (GROUP 2B) AUC = 0.974 [95% CI: 0.95, 1.00], **D** - COPD diagnosed by spirometry group (GROUP 3). AUC = 0.973 [95% CI: 0.95, 1.00].

Although the algorithm was developed to discriminate based on GOLD criteria we repeated the analysis using Lower Limit of Normal (LLN) thresholds to diagnose COPD. Test performance in the "COPD confirmed by spirometry group” (n=229) returned PPA of 100.0% [90.75%, 100.0%] and NPA of 75.4% [68.65%, 81.32%].

## DISCUSSION

We have described a simple, rapid diagnostic test for COPD which demonstrates high agreement with clinical diagnosis in the acute setting. Diagnostic agreement of the software algorithm with clinical diagnosis of COPD was PPA 93.8% and NPA 77.0%. Agreement was maintained when the patient had an acute respiratory infection (PPA 86.7% and NPA 80.5%). Notably, the index test retains high diagnostic agreement in cases of spirometry-confirmed COPD: PPA (100.0%) and NPA (77.0%).

Population and primary care surveys have demonstrated that mild (FEV_1_ ≥ 80% of percent predicted) and moderate (FEV^1^ 50-80% of percent predicted) airflow limitation is seldom diagnosed by clinicians [23, 24]. In our study, 48% of those with clinically-diagnosed COPD had only mild or moderate airflow limitation (Table 1). This group represents those who would benefit most from this algorithm, both by new treatment possibilities and also because they are frequently underdiagnosed.

We used the GOLD criteria for COPD diagnosis (FEV_1_/FVC<0.7) when developing our algorithms, although COPD can also be defined using the lower limit of normal (LLN). When calculated using the LLN thresholds, test performance was not significantly different from values obtained using GOLD criteria. It should be noted that as our model was developed to recognise COPD diagnosed using the GOLD criteria, we would expect a lower performance when the diagnostic criteria were changed.

In many European countries, spirometry is available in acute and primary care settings [8], however, uptake of the test is limited, leading to underdiagnosis or misdiagnosis of patients [6]. Several barriers to using spirometry in primary and acute care settings have been reported, including limitations in access, expertise and time; as well as expense [21]. Alternative testing methods have been developed. A meta-analysis of the COPD Diagnostic Questionnaire (CDQ) among ever smokers had a pooled sensitivity of 64.5% (95% CI 59.9% to 68.8%) and specificity 65.2% (52.9% to 75.8%) from four studies. Analysis of handheld flow meters showed a sensitivity of 79.9% (95% CI 74.2% to 84.7%) and specificity 84.4% (68.9% to 93.0%) from three studies [14]. In a scenario comparable to our study, when the CDQ was performed on symptomatic patients in primary care, the AUC was 0.65, sensitivity was 89.2%/65.8% and specificity 24.4%/54.0% for low risk and high risk of having COPD respectively [22]. The performance of our software algorithm exceeds that of currently available COPD screening questionnaires; outperforms the sensitivity of handheld flow meters with comparable specificity and demonstrates high agreement with the gold-standard (spirometry) in under one minute. This algorithm is intended to be used as a stand-alone device allowing for real-time diagnosis. As it is easy to operate and requires no physical patient contact, infection risk is minimised.

We envisage the algorithm could be used as an initial screening test in acute care settings for patients who present with non-specific respiratory symptoms. A positive result could be used to guide immediate care in the acute setting. As the test is delivered via smartphone, it could be applied in-person or during a telehealth consultation. A formal diagnosis of COPD requires confirmation by spirometry, the gold standard tool for COPD diagnosis [2]. Confirmatory spirometry could be performed during subsequent specialist follow up.

In this study, we were able to accurately identify the presence or absence of COPD in patients with LRTI, including pneumonia. In these situations, spirometry can be difficult to perform adequately, and an initial diagnostic test will help detect COPD in acutely unwell patients and identify those individuals most at risk of developing complications. Individuals with COPD are known to experience more frequent complications and mortality due to seasonal illnesses such as influenza [11]. More recently, a meta-analysis examining the risk of severe outcomes from SARS-CoV-2 infection (admission to ICU, mechanical ventilation or death) showed a greater than five-fold increase in risk of severe disease in patients with coexistent COPD [25]. We recommend that all COPD patients with a suspected infection should be carefully monitored in view of this increased risk. The diagnosis of COPD in patients presenting with SARS-CoV-2 or similar respiratory infections, would allow more focused therapeutic pathways and usefully guide healthcare resources to this at-risk group.

There are several limitations to this study. Our study population was recruited in an urban setting with smoking-related COPD. The generalisability of these results to COPD of differing aetiologies and in other settings requires confirmation. The tests were performed by trained research personnel in controlled environments, although we would consider the device less onerous to use than spirometry. The cough recording can be affected by background noise and positioning of the device, although the program will alert the user if background noise is excessive. The population recruited reflects the intended age range of use, however, as expected, those with diagnosed COPD were slightly older than those without and it will be important to replicate this study using an older control group.

The COPD diagnostic algorithm described in this study is used in combination with a suite of other respiratory diagnostic algorithms developed in the Breathe Easy program, including tests for asthma, pneumonia and lower respiratory tract disease [15]. The software provides a diagnostic output for each condition simultaneously every time it is used. Having independent decision algorithms for asthma, COPD and pneumonia is particularly important due to the considerable clinical and overlap between the conditions

In conclusion, the algorithm was able to accurately identify COPD even in the presence of infection. The algorithm operates as a stand-alone tool and provides a rapid result. It may find application in the acute-care setting as a screening tool to alert clinicians to the presence of COPD and allow more rapid, targeted and appropriate management.

## Data Availability

The underlying codes are the property of ResApp Health and are not available. The datasets supporting the conclusion of this article are available on reasonable request from PP and JB. The cough recordings are not available but will be uploaded as an educational tool in the future.

